# Human chromosomal-scale length variation and severity of COVID-19 infection using the UK Biobank dataset

**DOI:** 10.1101/2020.07.06.20147637

**Authors:** Chris Toh, James P. Brody

## Abstract

**Introduction:** The course of COVID-19 varies from asymptomatic to severe (acute respiratory distress, cytokine storms, and death) in patients. The basis for this range in symptoms is unknown. One possibility is that genetic variation is responsible for the highly variable response to infection. We evaluated how well a genetic risk score based on chromosome-scale length variation and machine learning classification algorithms could predict severity of response to SARS-CoV-2 infection.

**Methods:** We compared 981 patients from the UK Biobank dataset who had a severe reaction to SARS-COV-2 infection before 27 April 2020 to a similar number of age matched patients drawn for the general UK Biobank population. For each patient, we built a profile of 88 numbers characterizing the chromosome-scale length variability of their germ line DNA. Each number represented one quarter of the 22 autosomes. We used the machine learning algorithm XGBoost to build a classifier that could predict whether a person would have a severe reaction to Covid-19 based only on their 88-number classification.

**Results:** We found that the XGBoost classifier could differentiate between the two classes at a significant level *p* = 2 · 10 as measured against a randomized control and *p* = 3 · 10 measured against the expected value of a random guessing algorithm (AUC=0.5). However, we found that the AUC of the classifier was only 0.51, too low for a clinically useful test.

**Conclusion:** 

## Introduction

The course of COVID-19 varies from asymptomatic to severe (acute respiratory distress, cytokine storms, and death) in patients. The basis for this range in symptoms is unknown. One possibility is that genetic variation is responsible for the highly variable response to infection.

Human genetic variation can affect susceptibility and resistance to viral infections[1]. For instance, variants in the gene IFITM3 affect the severity of seasonal influenza[2]. Patients hospitalized from seasonal influenza had a particular allele of the gene IFITM3 at a higher rate than expected from the general population. Laboratory work determined that this particular allele can alter the course of the influenza virus infection.

We have previously shown that chromosome-scale length variation is a powerful tool to analyze genome wide associations[3]. This method is particularly appealing for genetic risk scores because it includes epistatic effects that might be missed with conventional genome wide association studies.

The purpose of this paper is to evaluate how well a genetic risk score based on chromosome-scale length variation and machine learning classification algorithms can predict severity of response to SARS-CoV-2 infection. We evaluated this approach on a dataset of 931 patients who had a severe reaction to Covid-19 in 2010. These patients had been previously genotyped as part of the UK Biobank.

## Methods

Data was obtained from the UK Biobank under Application Number 47850. First, we downloaded the “l2r” files from the UK Biobank. Each chromosome has a separate “l2r” file. Each “l2r” file contained 488,377 columns and a variable number of rows. Each column represented a unique patient in the dataset, who can be identified with an encoded ID number. Each row represented a different SNP. The values in the file represent the log base 2 ratio of intensity relative to the expected two copies measured at the SNP location.

After downloading the “l2r” data from the UK Biobank, we computed the mean l2r value for a portion of the chromosome for each patient in the dataset. This process produced a dataset where each person was represented by a series of 88 numbers. Each number represents the length variation for 25% of the 22 non-sex chromosomes. A value of 0 represents the nominal average length of that portion of the particular chromosome. We call this dataset the chromosome-scale length variation (CSLV) dataset.

This CSLV dataset was matched with the UK Biobank Covid-19 dataset. The Covid-19 data were provided to UK Biobank by Public Health England. UK Biobank matched the person in the Public Health England data with UK Biobanks internal records to produce the person’s encoded participant ID. The dataset we have, provided by UK Biobank contains the participant ID, date the specimen was taken, laboratory that processed the sample, whether the patient was an inpatient when the sample was taken, and the result (positive/negative) of the test. The UK Biobank continues to update the data approximately biweekly.

The criteria for testing and interpretation of results in the UK Biobank Covid-19 data has evolved. A positive test in this dataset earlier than 27 April 2020 was a good indication that the person had severe disease. During this early time period, SARS-CoV-2 testing was only performed on symptomatic people and this dataset only includes people tested in a hospital. After 27 April 2020, NHS instructed hospitals to test all non-elective patients admitted, including asymptomatic patients. The UK Biobank dataset released after 27 May 2020 includes “pillar 2” positive test results. These “pillar 2” tests include people in hospitals for non-elective procedures, staff screening, for care homes, and can include asymptomatic patients.

Using the CSLV-Covid-19 dataset, we selected all people who tested positive before 27 April 2020 and labelled these as people having a severe reaction to Covid-19. We segmented these into three overlapping datasets, as shown in Table 1. We constructed an age-matched control group of the same size that had an identical age profile as those in the severe reaction group. The age-matched control group was selected from the entire UK Biobank dataset, excepting those few who had a severe reaction to Covid-19. Since only a small fraction of the people in the UK Biobank had a severe reaction to Covid-19, we could rerun the analysis with a different age-matched control group many times to build up statistics. We chose this control group based on the data available and the finding that severe reactions to Covid-19 are a strong function of age and uncommon (only about 20% of those infected with SARS-CoV-2 require ICU admission even among those in their 70s)[4,5].

**Table 1.** We segmented the dataset into three overlapping subsets. The first, which we called “1930” contained all UK Biobank participants born after 1930 who had a severe reaction to SARS-CoV-2 infection before 27 April 2020. The two subsets contained people born after 1940 and after 1950.

| Dataset | Number |
| --- | --- |
| 1930 (< 90 years of age) | 981 |
| 1940 (< 80 years of age) | 880 |
| 1950 (<70 years of age) | 468 |

We used the H2O machine learning package in R to create XGBoost[6] models that were trained to classify a person in the dataset, consisting of those who had a severe reaction and age-matched controls, based solely on their chromosome scale length variation data.

## Results

The results are presented in Figure 1 and Table 2. As Figure 1 shows, we found a significant difference between all three age groupings and their corresponding random controls. This finding indicates that germ line genetics of the infected patient, as represented by the set of chromosome-scale length variation numbers, has an effect on the severity of COVID-19.

**Table 2.** We compared the difference in mean AUC values between the various datasets using a t-test. The datasets consisting of people born after 1930, 1940, and 1950 all showed significant differences with the corresponding random control. Those three datasets also showed significant differences between the mean AUC and 0.5. The three random controls did not show a significant difference between the mean AUC and 0.5, as expected. An AUC value of 0.5 represents a random classification test, one in which the algorithm is no better than guessing.

|  |  | **p-value of t-test** |
| --- | --- | --- |
| 1930 data | 1930 random | $2 \cdot 10^{-11}$ |
| 1940 data | 1940 random | $1 \cdot 10^{-9}$ |
| 1950 data | 1950 random | $1 \cdot 10^{-4}$ |
| 0.5 | 1930 data | $3 \cdot 10^{-14}$ |
| 0.5 | 1940 data | $4 \cdot 10^{-13}$ |
| 0.5 | 1950 data | $3 \cdot 10^{-4}$ |
| 0.5 | 1930 random | 0.1 |
| 0.5 | 1940 random | 0.4 |
| 0.5 | 1950 random | 0.08 |

**Figure 1.**
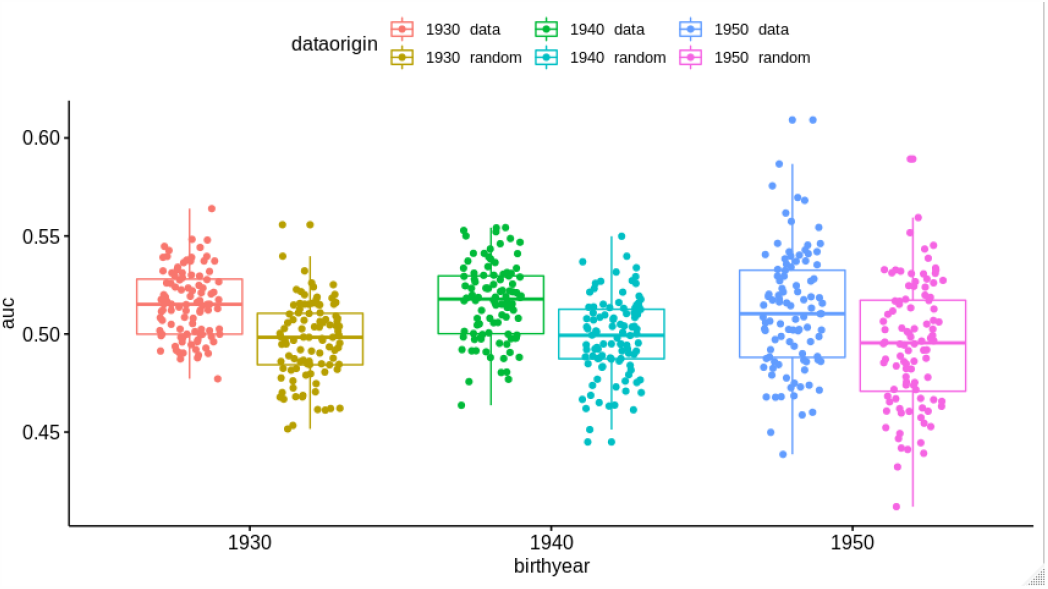
This boxplot figure presents the results of the machine learning predictions. We created three different datasets, one which includes all patients less than 90 years old, the second includes every patient less than 80 years old, and the third with every patient less than 70 years old. These are indicated as the oldest birthyear “data”. Each dataset included an equal number of patients with a “severe reaction” to Covid-19 and an equal number of age matched people drawn from the general UK Biobank population, “normal”. For comparison, we took those three datasets and randomly permuted the status (“severe reaction” or “normal”) and repeated the process. This randomly permuted dataset is labelled oldest birthyear “random”. For each dataset, we repeated the whole process 100 times, each time with a different set of age matched people from the general UK Biobank population.

Figure 1 and Table 3 also show that the AUC (area under the curve of the receiver operating characteristic curve) for the XGBoost classification model was about 0.51. A classification model with an AUC of 0.51 is just slightly better than guessing.

**Table 3.** The mean, and standard deviation, of the area under the curve of the receiver operating characteristic curve was recorded after 100 different runs of the XGBoost model. Each run used a different set of people who did not have a severe reaction to Covid-19. The mean AUC for all three datasets was well described by a normal distribution, as confirmed by a Shapiro normality test.

|  | **Mean AUC** | **SD AUC** |
| --- | --- | --- |
| 1930 data | 0.515 | 0.017 |
| 1940 data | 0.516 | 0.019 |
| 1950 data | 0.511 | 0.030 |

## Discussion

The two conclusions of this study are divergent. First, a genetic difference exists between those who have the most severe course of the disease and the general population. Second, we were not able to exploit this difference to develop a clinically useful test to distinguish between people who will experience a severe course of the disease and those who will not. We could only demonstrate a genetic risk test with an AUC of 0.51, only slightly above 0.50 which represents random guessing.

Although the AUC we found here is too low to be clinically useful, several avenues for improving the AUC exist. We were constrained by the data available to compare those who had a severe reaction to Covid-19 with the general population, but the general population probably contains a substantial number of people who would also have a severe reaction to Covid-19. A better approach would be to compare those who had a severe reaction to Covid-19 with those who were asymptomatic or had a mild reaction. Simply having more data on those patients who had a severe reaction might also lead to an increase in AUC. We could also have more data on each individual patient. The algorithm we used for transforming “l2r” data into our final chromosome-scale length variation data took averages over each quarter of a chromosome. We could instead include smaller chromosome segments. Finally, an alternative machine learning algorithm might provide improved AUC. Different algorithms perform differently on different classes of problems. [7] We did a brief test of different algorithms before choosing XGBoost for this problem, but, for instance, a deep learning algorithm might have superior performance with proper tuning.

Our results add to the recent work done by other on the link between genetics and severity of Covid-19. A detailed study of this UK Biobank Covid-19 dataset identified that Black and Asian patients were at a significantly higher risk of testing positive compared to white patients [8]. This study also attempted to derive a polygenic risk score. However, when they applied the polygenic risk score to a hold-out group, they found that the mean score was indistinguishable between the group of people who had tested positive and the group that had no positive test. In comparison, our work found that these two groups are distinguishable with a genetic risk score, but only very slightly. We measured the AUC at 0.51. They do not report an AUC, but an indistinguishable test is the equivalent of an AUC of 0.50.

Other more comprehensive metastudies have identified one specific genetic component behind the severity of Covid-19. For instance, one study of Covid-19 patients who experienced respiratory failure at seven hospitals in Italy and Spain found a fairly strong association in a cluster of genes lying on part of chromosome 3 and a borderline association in chromosome 9 encompassing the ABO blood group locus [9]. The June 2020 results posted by the Covid-19 Host Genetics Initiative [10,11], also indicate a strong association in Chromosome 3, but fail to reproduce the association in chromosome 9. The Covid-19 Host Genetics Initiative “ANA_B2” study represents hospitalized Covid-19 patients compared to the general population and are derived from mostly patients in Europe and Brazil. Neither study attempted to derive a genetic risk score.

## Conclusion

In conclusion, we found a significant difference exists between the structural genomics of those patients in the UK Biobank who had a severe reaction to the SARS-CoV-2 virus and the general UK Biobank population. However, a test based upon this difference would not be clinically useful in its present state, since it had an AUC of 0.51.

## Data Availability

The original data can be obtained from the UK Biobank.

https://www.ukbiobank.ac.uk/

## Acknowledgements

The data used in this study was obtained from the UK Biobank under Application Number 47850.

## Notes

### Competing Interest Statement

The authors have declared no competing interest.

### Funding Statement

No external funding was received for this work.

### Author Declarations

This data uses previously collected data from UK Biobank. The original consent was obtained by UK Biobank. Our analysis of the de-identified data has been examined by UC Irvine's IRB. They ruled that our research is not considered "human subjects research" because it uses de-identified data.

## References

1. Kenney AD, Dowdle JA, Bozzacco L, McMichael TM, St Gelais C, Panfil AR, et al. Human Genetic Determinants of Viral Diseases. Annual review of genetics. 2017;51: 241–263. doi:10.1146/annurev-genet-120116-023425

2. Everitt AR, Clare S, Pertel T, John SP, Wash RS, Smith SE, et al. IFITM3 restricts the morbidity and mortality associated with influenza. Nature. 2012;484: 519–23. doi:10.1038/nature10921

3. Toh C, Brody JP. Analysis of copy number variation from germline DNA can predict individual cancer risk. bioRxiv. 2018; 303339. doi:10.1101/303339

4. Davies NG, Klepac P, Liu Y, Prem K, Jit M, Eggo RM. Age-dependent effects in the transmission and control of COVID-19 epidemics. Nature Medicine. 2020; 1–7. doi:10.1038/s41591-020-0962-9

5. Bialek S, Boundy E, Bowen V, Chow N, Cohn A, Dowling N, et al. Severe Outcomes Among Patients with Coronavirus Disease 2019 (COVID-19) — United States, February 12–March 16, 2020. MMWR Morbidity and Mortality Weekly Report. 2020;69: 343–346. doi:10.15585/mmwr.mm6912e2

6. Chen T, Guestrin C. XGBoost: A Scalable Tree Boosting System. 2016 [cited 9 May 2018]. doi:10.1145/2939672.2939785

7. Olson RS, Cava W la, Mustahsan Z, Varik A, Moore JH. Data-driven advice for applying machine learning to bioinformatics problems. Pacific Symposium on Biocomputing Pacific Symposium on Biocomputing. 2018;23: 192–203. Available: http://www.ncbi.nlm.nih.gov/pubmed/29218881

8. Kolin DA, Kulm S, Elemento O. Clinical and Genetic Characteristics of Covid-19 Patients from UK Biobank. medRxiv. 2020; 2020.05.05.20075507. doi:10.1101/2020.05.05.20075507

9. Ellinghaus D, Degenhardt F, Bujanda L, Buti M, Albillos A, Invernizzi P, et al. Genomewide Association Study of Severe Covid-19 with Respiratory Failure. New England Journal of Medicine. 2020; NEJMoa2020283. doi:10.1056/NEJMoa2020283

10. The COVID-19 Host Genetics Initiative, a global initiative to elucidate the role of host genetic factors in susceptibility and severity of the SARS-CoV-2 virus pandemic. European Journal of Human Genetics. 2020;28: 715–718. doi:10.1038/s41431-020-0636-6

11. Covid-19 Host Genetics Initiative Results. [cited 29 Jun 2020]. Available: https://www.covid19hg.org/results/

